# spread.gl: visualising pathogen dispersal in a high-performance browser application

**DOI:** 10.1101/2024.06.04.24308447

**Authors:** Yimin Li, Nena Bollen, Samuel L. Hong, Marius Brusselmans, Fabiana Gámbaro, Marc A. Suchard, Andrew Rambaut, Philippe Lemey, Simon Dellicour, Guy Baele

## Abstract

Phylogeographic analyses are able to exploit the location data associated with sampled molecular sequences to reconstruct the spatio-temporal dispersal history of a pathogen. Visualisation software is commonly used to facilitate the interpretation of the accompanying estimation results, as these are not always easily interpretable. spread.gl is a powerful, open-source and feature-rich browser application that enables smooth, intuitive and user-friendly visualisation of both discrete and continuous phylogeographic inference results, enabling the animation of pathogen geographic dispersal through time. spread.gl can render and combine the visualisation of several data layers, including a geographic layer (e.g., a world map), multiple layers that contain information extracted from the input phylogeny, and different types of layers that represent environmental data. As such, users can explore which environmental data may have shaped pathogen dispersal patterns, that can subsequently be formally tested through more principled statistical analyses. We showcase the visualisation features of spread.gl on several representative pathogen dispersal examples, including the smooth animation of a phylogeny encompassing over 17,000 genomic sequences resulting from a large-scale SARS-CoV-2 analysis.

## Introduction

Statistical phylogeographic inference has become widely adopted as a method to investigate viral circulation, often offering the ability to reconstruct the migration history of pathogens in a spatial-temporal context. Phylogeographic models and methods have been implemented in various software packages, each employing its own inference procedures. BEAST 1.10 (Suchard et al., 2018) and BEAST 2 (Bouckaert et al., 2019) are widely-used software packages for Bayesian phylogeographic analysis, whereas TreeTime (Sagulenko et al., 2018) and PastML (Ishikawa et al., 2019) employ maximum-likelihood inference to infer ancestral discrete locations. While the use of discrete phylogeographic inference may be more widespread than its continuous counterpart, the applicability and performance of both approaches remain a topic of continued investigation (Kalkauskas et al., 2021; Layan et al., 2023), which will undoubtedly lead to further improvements and novel implementations.

To visualise and interpret the outcomes of phylogeographic analyses, a range of software packages have been developed to parse location-annotated phylogenies, which contain spatial and temporal information, making them well-suited to create animations through time. Arguably, the most popular (online) visualisation software package resource in recent years is Nextstrain (Hadfield et al., 2018). Auspice is an open-source interactive tool that powers the visualisations within Nextstrain, allowing customisation of aesthetics and functionality, making use of associated metadata such as geographic information, serology, or host species. Nextstrain offers a visual interface that consists of three linked panels: a phylogenetic tree, the inferred transition events among locations, and the genetic diversity across the pathogen’s genome. PastML (Ishikawa et al., 2019) enables a visualisation through a zoomable HTML map while performing a two-step compression: a vertical compression clusters together the regions of the tree where no state changes occur, whereas a horizontal compression merges identical subtree configurations. SPREAD (Bielejec et al., 2011), SpreaD3 (Bielejec et al., 2016), and SPREAD4 (Nahata et al., 2022) are a set of successive improvements of software used to analyse and visualise the results of phylogeographic reconstructions. A short overview of other visualisation applications is discussed in the recent SPREAD4 publication (Nahata et al., 2022). Certain visualisation approaches, such as the one implemented in the R package “seraphim” (Dellicour et al., 2016) or the Python library “baltic” (https://github.com/evogytis/baltic), offer great flexibility but require programming knowledge to visualise a pathogen’s dispersal history.

Each of the tools mentioned above has its own advantages and disadvantages, the latter of which may be inflexible customisation, unintuitive user interfaces, as well as compatibility and dependency issues. For example, SPREAD relied on the Google Earth software and as such required converting a consensus tree or posterior set of trees to Keyhole Markup Language (KML). Its successor, SpreaD3, no longer relied on external software nor on having an internet connection but became sensitive to updates to many of its supported browser platforms. SPREAD4, a cloud-based successor of SpreaD3, currently employs Amazon Web Services for its calculations and features account-based storage and easy sharing of visualisations by means of unique web addresses. However, it is still inconvenient for users to adjust elements of its phylogeographic animations, such as the start and end times, the current time point, and the playback speed. Furthermore, a number of visualisation packages focus on a specific type of phylogenetic inference (i.e., discrete or continuous), and may hence not offer support for both types. Apart from these issues, most applications struggle to visualise phylogeographic reconstructions of large data, an issue that became all too apparent when studying the spread of SARS-CoV-2 lineages. When a large phylogenetic tree is loaded, a range of potential issues can emerge, such as long processing times, unsatisfactory responsiveness and obvious animation stuttering.

To address the various limitations mentioned above, we have developed spread.gl, a high-performance open-source browser-based visualisation application. This latest instalment in the continuing SPREAD development focuses on smooth and high-performance animations, improved aesthetics and extensive functionality. A key focus of this new version is to enable the simultaneous visualisation of many data layers, with a typical use case being the inclusion of environmental data layers upon which a pathogen’s dispersal through time and space can be visualised. Further, the order of these different types of layers is made adjustable and can be easily enabled or disabled. We employ these various (types of) data layers to incorporate environmental and ecological factors that offer potentially important context for the dispersal history and spatio-temporal patterns of pathogens, which is not possible in the other software packages mentioned above. We showcase the capabilities of spread.gl through pathogen dispersal examples in the *Visualisation examples* section.

### Software features

To accomplish the aforementioned functionalities, spread.gl was built upon kepler.gl – a powerful open-source framework for geospatial analysis of big data – to enable the efficient rendering of large-scale phylogenies on maps with highly customisable visualisation options. kepler.gl – and by extension spread.gl – is a client-side-only application, meaning that the data you upload to the application are only stored in the browser and no information or map data is sent to any server, which is of particular interest when working with health data or any type of data with privacy concerns. Three different data formats are accepted as input: comma-separated values (CSV), JavaScript object notation (JSON) and GeoJSON, an open standard format designed for representing geographical features, along with their non-spatial attributes. Phylogenetic trees output from Bayesian phylogeographic analyses are typically stored as NEXUS files so we provide a way to convert them into an acceptable input data format. A key step consists of creating an *arc layer* to display the phylogenetic branches, combined with additional layers to visualise important elements related to the ancestral location nodes from discrete or continuous phylogeographic inference.

An important addition in spread.gl, compared to previous versions of SPREAD, is the ability to visualise phylogeographic reconstructions on top of an environmental data layer to enable a visual exploration of possible associations with pathogen dispersal. Construction of an environmental layer is relatively straightforward (see the online tutorial mentioned in the *Software availability* section), and several such layers can be combined within a single visualisation. Once all the layers of interest have been prepared and added, an animation of pathogen dispersal can be generated and played over an incremental or moving time window.

When a visualisation has been constructed to the user’s satisfaction, the final result can be exported as an image or a JSON file, an HTML file – which can easily be shared with others who are not required to have spread.gl installed – or captured from the screen using a video capture tool. The final result of spread.gl can also be exported as a PDF using a browser extension such as GoFullPage (Coles, 2023), which is free to use and compatible with different browsers, e.g. Google Chrome and Microsoft Edge. Safari on MacOS allows to directly save the web page as a PDF.

In the examples below, we focus on parsing the output from the BEAST 1.10 (Suchard et al., 2018) and BEAST 2 (Bouckaert et al., 2019) software packages, arguably the most widely used tools for studying a pathogen’s spatio-temporal dispersal patterns. The output of other software packages can of course also be used, as long as the output format is the same, or is able to be converted using one of the corresponding processing scripts (see the *Software availability* section).

#### Visualisation of discrete phylogeographic inference

When visualising the outcome of a discrete phylogeographic analysis, in addition to displaying the phylogenetic branches using an *arc layer*, one can create an additional *cluster layer* to emphasise the relevance of each ancestral location in the pathogen’s dispersal history. Such a layer attaches a circle centred on each ancestral location, with a radius proportional to the number of lineages present in that location. This cluster layer not only visually represents the varying intensity at each location of pathogen spread based on the accumulated ancestral lineages in a specific period, but also provides additional details of absolute counts by hovering the mouse over the circles. We also provide a Bayes factor test to identify well-supported rates between locations by estimating their statistical support through Bayesian stochastic search variable selection (BSSVS) (Lemey et al., 2009). Once the Bayes factors have been computed (see the online tutorial link in *Software availability*), they can be added as a filter in spread.gl so as to only display the transition rates that are well-supported by the data.

#### Visualisation of continuous phylogeographic inference

When visualising the outcome of continuous phylogeographic inference, the *arc layer* that represents the phylogenetic branches can be combined with a *point layer* to show the sampled genomes as well as their ancestral locations. Furthermore, a *contour layer* comprised of semi-transparent polygons can be created to depict the highest posterior density (HPD) region for each ancestral node location and to visually represent the geographic uncertainty of the location estimates (Lemey et al., 2010).

#### Visualisation of large-scale phylogeographic inference

Large-scale phylogenetic and phylogeographic analyses present a myriad of potential processing problems, including how to perform (any type of) Bayesian phylogeographic inference and how to generate maximum clade credibility (MCC) trees from the large output files that contain annotations for thousands or tens of thousands of ancestral nodes. As such, the output phylogeny from such large-scale studies may – by necessity – be a single (e.g., the final) location-annotated phylogeny sampled from the posterior distribution (see e.g. Kraemer et al. (2021)) without any uncertainty estimates for the estimated locations at the internal nodes. During the visualisation process, such a single tree may not be easily processed as it typically stores the exact geographical coordinates of each location within a single annotation, rather than as two distinct continuous traits in a standard MCC tree. Additionally, the use of various coordinate reference systems (CRS) necessitates specific programming expertise in geographic coordinate conversion, which can be challenging for users to implement proper re-projection on the map. spread.gl is designed to address the challenges described in these scenarios, enabling the smooth and responsive visualisation and animation of large data.

#### Visualisation of environmental layers

When studying the spread and evolution of pathogens, it is often of interest to investigate which environmental factors may potentially influence their dispersal. To facilitate such a visual exploration in spread.gl, we provide the opportunity to overlay environmental and/or ecological layers. Such data are often available in the public domain and can be freely downloaded from online resources. There are two primary types of these data, i.e. raster and vector data, both of which are supported in the spread.gl toolkit. Raster data come in the form of a geo-referenced matrix where each cell represents a geographic region and contains an attribute value that measures a particular characteristic of that region. This measurement can be discrete, to represent distinct categories such as land cover variables, or continuous, to store gradually changing values such as temperature and elevation. Vector data consist of different geometric features and multiple assigned attributes that store additional information, with GeoJSON being one of the most general vector data formats, which is supported in spread.gl. To assign such data to the locations of interest, the actual shapes of the locations of interest (e.g. countries, provinces, etc.) need to be retrieved. Shape data are also typically freely available online either in a shape file or in the GeoJSON format. Both a *point layer* and a *polygon layer* can be used in spread.gl to visualise these additional layers. For example, a *point layer* – with a customisable radius of the points – can be used to show gradually changing environmental data, whereas a *polygon layer* is typically used to show values – e.g. the gross domestic product (GDP) – for larger administrative areas.

### Visualisation examples

In the sections below, we introduce three examples of phylogeographic studies and showcase the visualisation of the inference results in spread.gl. These constitute the primary use cases for spread.gl but there are many more custom visualisations that can be created (see the GitHub repository and the provided example HTML output files as mentioned in the *Software availability* section).

#### SARS-CoV-2 lineage B.1.1.7 in England (large-scale continuous phylogeography)

In a study conducted by Kraemer et al. (2021), the dispersal history and dynamics of the B.1.1.7 lineage of severe acute respiratory syndrome coronavirus 2 (SARS-CoV-2) were analysed using a continuous phylogeography approach based on 17,716 genomes. This particular variant was initially identified in Kent or Greater London in September 2020 and rapidly spread throughout the United Kingdom by the end of that year, eventually becoming a variant of global concern. To visualise the continuous phylogeographic reconstruction from this study in spread.gl, we have performed the following steps. First, to achieve the correct projection, we re-projected the original coordinates by converting the CRS from the British National Grid (BNG) to the current World Geodetic System (WGS84) (to this end, a script is available in the GitHub repository of spread.gl). As a second step, we meticulously identified and removed outliers, which were data points that appeared outside England in the initial visualisation, as these points lacked reliable geospatial information in the metadata. As a result, the total number of visualised genomes was reduced to 17,203. Finally, we created a *point layer* and an *arc layer* to represent the spatio-temporal information in the location-annotated phylogeny, for which we took the final sample from the original Bayesian continuous phylogeographic analysis. The end result is displayed in Figure 1. By smoothly rendering an animated display of the many sampling points and phylogenetic branches, spread.gl showcases its high performance and robust support for handling large-scale datasets.

**Fig. 1.**
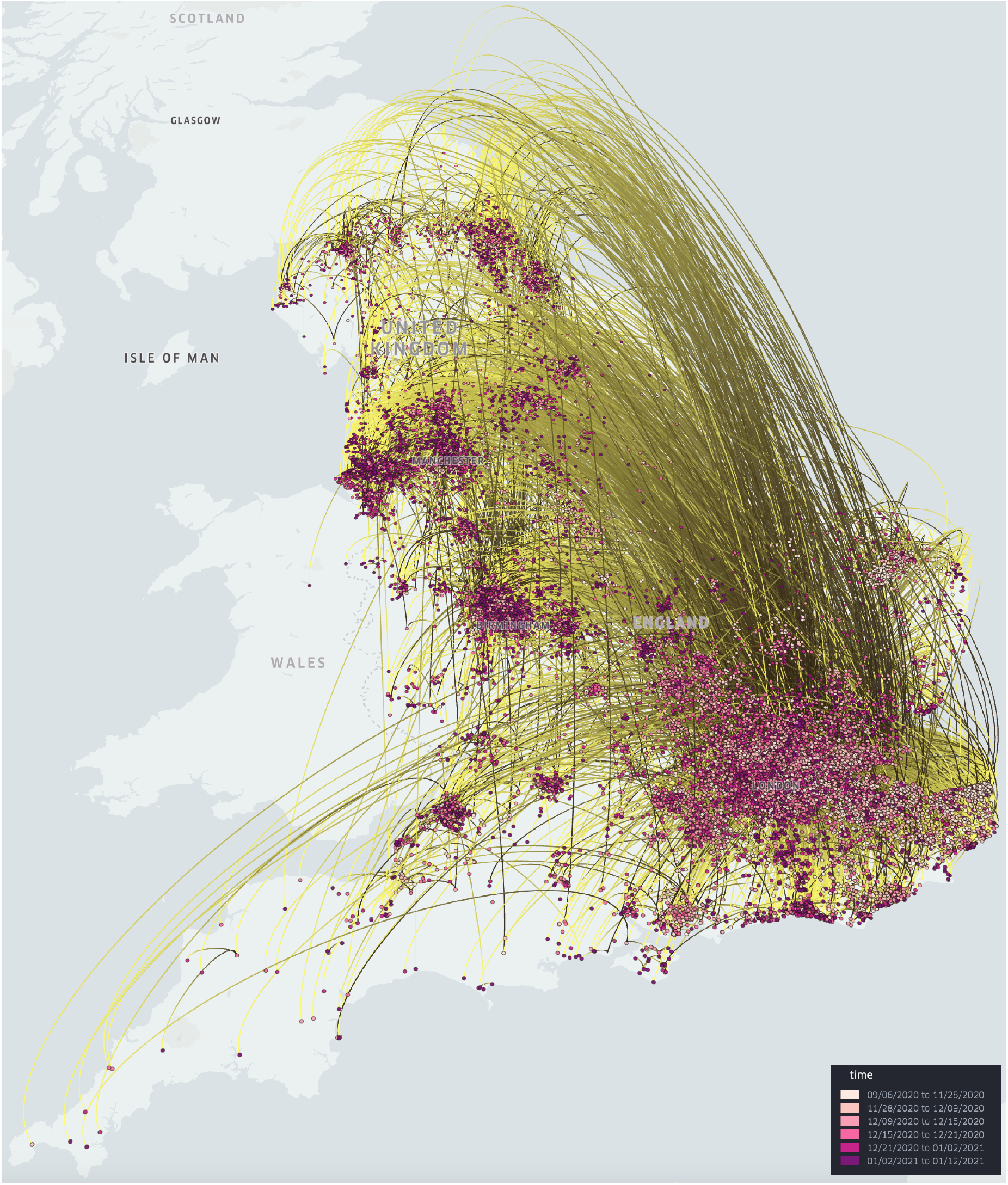
Continuous phylogeographic reconstruction of the dispersal history of SARS-CoV-2 lineage B.1.1.7 in England (in 3D mode). This visualisation of a large SARS-CoV-2 dataset containing 17,203 genomes can be rendered efficiently using spread.gl, with animation of the spread over time being shown smoothly despite a large number of branches in the phylogeny.

#### YFV in Brazil (continuous phylogeography with environmental data)

Hill et al. (2022) conducted an analysis of the outbreak of yellow fever virus (YFV) in Brazil between 2016 and 2019. The author reconstructed the spatio-temporal spread based on 705 YFV sequences, generated from neotropical primate, human and mosquito samples. This study considered the effects of environmental factors on the spread of YFV and identified a significant association between temperature and virus effective population sizes. We downloaded historical maximum temperature data from WorldClim (Fick and Hijmans, 2017; Harris et al., 2020) and created a *point layer* of the average maximum temperatures across the area that covers the relevant Brazilian states during the virus outbreak. To balance file size and granularity, temperature data with a spatial resolution of 2.5 minutes (∼21 km^2^ at the equator) were used in our visualisation. The resulting raster file was clipped using a mask layer consisting of a boundary map and a list of locations, to display only the Brazilian states of interest in the spread.gl visualisation. On top of this environmental layer, we created a *point layer*, an *arc layer* and a *contour layer* to represent the spatio-temporal information (and its associated uncertainty) in the location-annotated MCC tree.

In Figure 2, we show the complete spread.gl visualisation of the continuous phylogeographic reconstruction on top of an environmental data layer representing the global maximum temperature averaged over all months during the outbreak, which corresponds to an environmental factor suspected to impact the genetic diversity and dispersal dynamics of the virus (Hill et al., 2022). Improving the transparency of the contour layer, achieved by reducing the opacity of the polygons from a standard 20% to a lower 5%, ensures that the temperature layer is not excessively obscured to facilitate a clearer understanding of the dynamics between temperature and state transitions.

**Fig. 2.**
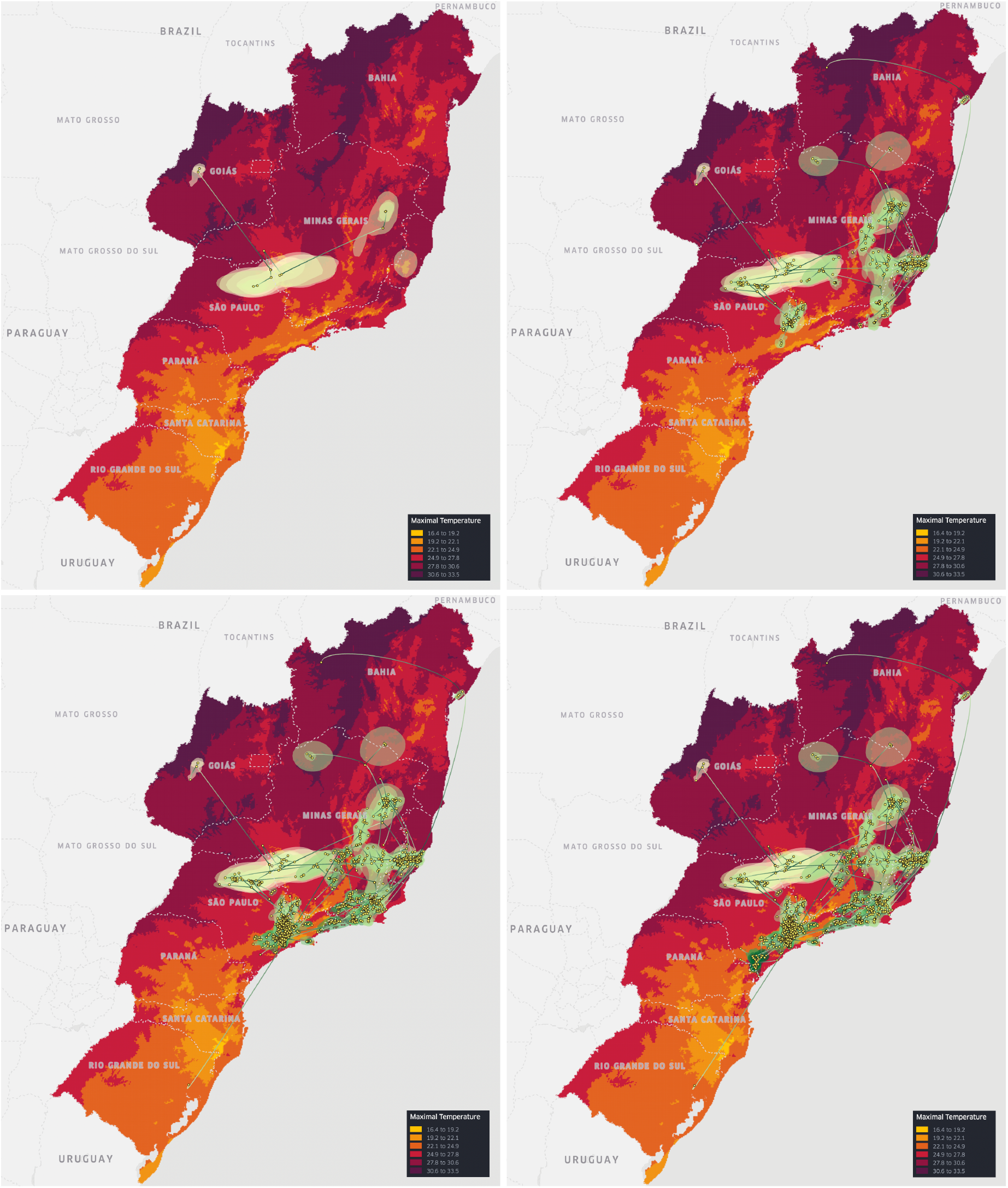
Continuous phylogeographic reconstruction of the dispersal history of yellow fever virus lineages in Brazil (in 2D mode) on top of a (maximal) temperature data layer: a) until April 7, 2016; b) until April 10, 2017; c) until April 12, 2018; d) until April 16, 2019.

#### PEDV in China (discrete phylogeography with environmental data)

Porcine epidemic diarrhoea virus (PEDV) generally results in high mortality in infected suckling pigs. To study the dispersal pattern of this virus, He et al. (2021) conducted a discrete phylogeographic analysis of a data set that contained 769 sequences sampled in 26 provinces across China and unravelled the spread of PEDV genotype G2 during the past two decades. In their study, the authors explored a range of environmental factors of interest to determine which of them had an important impact on the dispersal of PEDV between provinces in China. We first curated an environmental layer as a *Geojson layer* in spread.gl, containing 16 environmental variables of interest related to economy, demography and geography for each province: sample size, slaughtered pigs, pork price, live pig price, pork consumption, total consumption, feed price, feed production, GDP, human population size, human population density, maximum elevation, mean elevation, mean temperature, mean vapour pressure, and mean precipitation. As shown in Figure 3, each province can be coloured based on the corresponding value of an environmental variable, in this case, mean pork consumption per capita (kg) between 2015 and 2018. On top of this environmental layer, we displayed an *arc layer* to show the phylogenetic branches and a *cluster layer* to reflect the relevance of each ancestral location in the pathogen’s dispersal. Based on an intuitive visual assessment, pork consumption does not show a pronounced impact on the spread of PEDV in China, although it was described as relevant to and indirectly associated with viral transmission in the original study (He et al., 2021). Indeed, from the environmental layer in Figure 3 we observe that most of the provinces in South China have high pork consumption. However, from the reconstruction of PEDV viral spread, we observe that both Guangdong and Henan (i.e., not provinces with high pork consumption) are the main hubs for the spread of PEDV within China, followed by Sichuan. These observations point to the importance of combining visual inspections with detailed statistical assessments to determine the impact of predictors on viral spread.

**Fig. 3.**
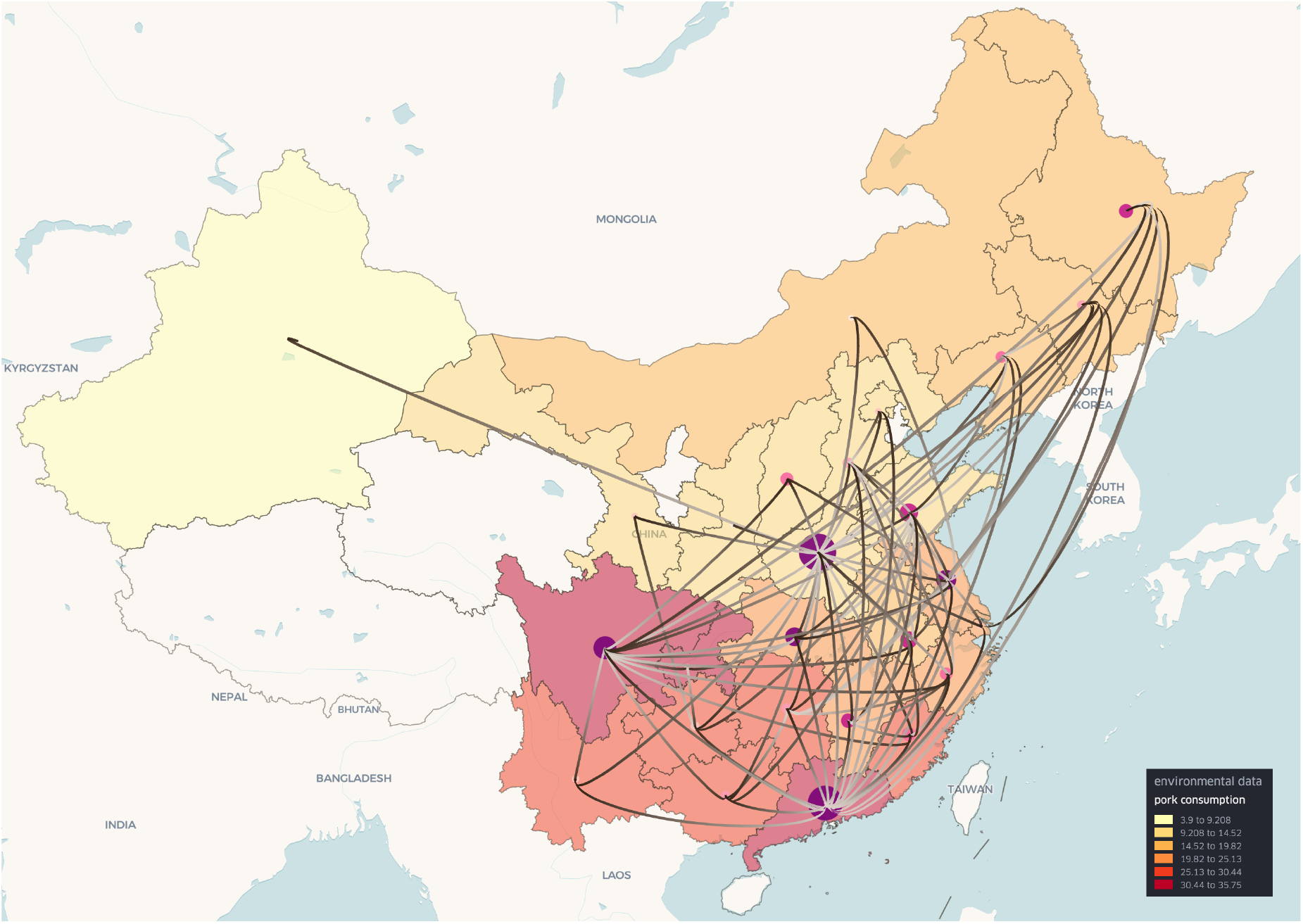
Discrete phylogeographic reconstruction of the spread of porcine epidemic diarrhoea virus in China (in 2D mode) on top of a semi-transparent data layer that shows pork consumption per province. Provinces for which these data are not available are not visualised. Colours towards the yellow spectrum indicate lower pork consumption, whereas colours towards the (dark) red spectrum indicate higher pork consumption. The start and end of the branches are coloured in white and grey, respectively, with the colour gradually changing across the length of the branches. Darker purple clusters represent locally higher cumulative lineage counts over a custom period, while lighter pink clusters represent lower cumulative counts.

## Data Availability

All the required source code and installation instructions for spread.gl can be found on GitHub, as well as video animations of the visualisations presented in this article: https://github.com/GuyBaele/SpreadGL.
HTML output from the spread.gl visualisations shown in this article can be readily downloaded from: https://github.com/GuyBaele/SpreadGL/tree/main/html.
All the required input data and processing scripts - along with instructions on how to use them - for the spread.gl visualisations shown in this article can be found on: https://github.com/GuyBaele/SpreadGL/tree/main/inputdata.

## Software availability

All the required source code and installation instructions for spread.gl can be found on GitHub, as well as video animations of the visualisations presented in this article: https://github.com/GuyBaele/SpreadGL. HTML output from the spread.gl visualisations shown in this article can be readily downloaded from: https://github.com/GuyBaele/SpreadGL/tree/main/html.

All the required input data and processing scripts – along with instructions on how to use them – for the spread.gl visualisations shown in this article can be found on: https://github.com/GuyBaele/SpreadGL/tree/main/inputdata.

## Author contributions statement

G.B. conceived the study. Y.L. designed and developed the software. Y.L., N.B. and S.L.H. collected the data. G.B., Y.L., N.B. and S.D. analysed the results. G.B., Y.L., N.B., S.L.H., M.B., F.G. and S.D. tested the software. G.B., Y.L. and N.B. wrote and revised the manuscript. S.D., S.L.H., P.L., M.A.S. and A.R. reviewed the manuscript. G.B. and S.D. acquired funding for the study.

## Acknowledgments

Y.L., N.B., S.D. and G.B. acknowledge support from the Research Foundation - Flanders (“Fonds voor Wetenschappelijk Onderzoek - Vlaanderen,” G098321N). S.L.H. and G.B. acknowledge support from the Research Foundation - Flanders (“Fonds voor Wetenschappelijk Onderzoek - Vlaanderen,” G0E1420N). S.D. and G.B. acknowledge support from the European Union Horizon 2023 RIA project LEAPS (grant agreement no. 101094685). S.D. acknowledges support from the “Fonds National de la Recherche Scientifique” (F.R.S.-FNRS, Belgium; grant no. F.4515.22). S.D. and P.L. acknowledge support from the European Union Horizon 2020 project MOOD (grant agreement no. 874850). G.B. acknowledges support from the Internal Funds KU Leuven (Grant No. C14/18/094). P.L., M.A.S. and A.R. acknowledge support from the Wellcome Trust (Collaborators Award 206298/Z/17/Z, ARTIC network), the European Research Council (grant agreement no. 725422 – ReservoirDOCS) and the National Institutes of Health (R01 AI153044). P.L. acknowledges support from the Research Foundation, Flanders (“Fonds voor Wetenschappelijk Onderzoek - Vlaanderen,” G066215N, G0D5117N and G0B9317N). G.B. and M.B. acknowledge support from the DURABLE EU4Health project 02/2023-01/2027, which is co-funded by the European Union (call EU4H-2021-PJ4) under Grant Agreement No. 101102733. Views and opinions expressed are however those of the author(s) only and do not necessarily reflect those of the European Union or the European Health and Digital Executive Agency. Neither the European Union nor the granting authority can be held responsible for them.

---

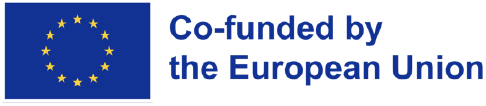

